# Bridging the translational neuroscience gap: Development of the ‘shiftability’ paradigm and an exemplar protocol to capture psilocybin-elicited ‘shift’ in neurobiological mechanisms in autism

**DOI:** 10.1101/2023.05.25.23290521

**Authors:** Tobias P. Whelan, Eileen Daly, Nicolaas A. Puts, Ekaterina Malievskaia, Declan G.M. Murphy, Grainne M. McAlonan

**Affiliations:** Department of Forensic and Neurodevelopmental Sciences, Institute of Psychiatry, Psychology & Neuroscience, King’s College London, UK; COMPASS Pathfinder Ltd, London, UK; Sackler Institute for Translational Neurodevelopment, Institute of Psychiatry, Psychology & Neuroscience, King’s College London, UK; MRC Centre for Neurodevelopmental Disorders, King’s College London, UK

## Abstract

Clinical trials of pharmacological approaches targeting the core features of autism have failed. This is despite evidence from preclinical studies, genetics, post-mortem studies and correlational analyses linking peripheral and central markers of multiple candidate neurochemical systems to brain function in autism. Whilst this has in part been explained by the heterogeneity of the autistic population, the field has largely relied upon association studies to link brain chemistry to function. The only way to *directly* establish that a neurotransmitter or neuromodulator is involved in a candidate brain function is to *change* it and observe a *shift* in that function. This experimental approach dominates preclinical neuroscience, but not human studies. There is very little direct experimental evidence describing how neurochemical systems modulate information processing in the living human brain. As a result, our understanding of how neurochemical differences contribute to neurodiversity is limited and impedes our ability to translate findings from animal studies into humans.

Here, we begin by introducing our “shiftability” paradigm, an approach to bridge the translational gap in autism research. We then provide an overview of the methodologies used and explain our most recent choice of psilocybin as a pharmacological probe of the serotonin system *in vivo*. Finally, we provide a summary of the protocol for ‘PSILAUT’, an exemplar “shiftability” study which uses psilocybin to directly test the hypothesis that the serotonin system functions differently in autistic and non-autistic adults.

## 1. Introduction

Autistic spectrum disorder (ASD), the overarching definition used by our current diagnostic guidelines, is a life-long neurodevelopmental condition characterised by differences in social interaction and communication, repetitive or restricted patterns of behaviour, and sensory differences^1^. Clinical trials have failed to offer pharmacological options for those that seek support. This in part reflects the complex neurobiology and diversity of autism. However, clinical trial efforts have also been hampered because there is no reliable experimental bridge from ‘basic’ neuroscience to clinical application. This is not an isolated problem; it is common across neuropsychiatric conditions. We do not yet fully understand how neurochemical systems regulate information processing within and across different levels of brain organisation in neurotypical or neurodivergent people, and in particular, how this links to core traits of these conditions.

A constraint is that human neuroscience research has largely relied on association or correlational (and cross-sectional) evidence linking genetic, preclinical, post-mortem and neuroimaging data to, for example, trait(s), symptom(s) or diagnoses. However, correlations are not causal. The only way to establish that a neurosignalling system is involved in a candidate brain mechanism is to change it and observe a ‘shift’ in that mechanism. This is basis of preclinical study designs, but there has been limited translation to humans.

We have developed a direct experimental approach to examine neurochemical regulation of information processing across the organisational levels of the human brain which is sensitive to *individual* differences in neurotypical and neurodivergent people. We call this a “shiftability” paradigm. To date, our “shiftability” studies have demonstrated that the autistic brain responds differently when different neurochemical systems are perturbed by a single, low dose pharmacological challenge^2–11^.

Here, we begin by introducing our “shiftability” paradigm, a novel approach to bridge the translational gap in autism research. We then provide an overview of the methodologies used and explain our most recent choice of psilocybin as a pharmacological probe of the serotonin system *in vivo*. Finally, we provide a summary of the protocol for ‘PSILAUT’, our latest “shiftability” study which uses psilocybin to directly test the hypothesis that the brain’s serotonin system functions differently in autistic and non-autistic adults. The end goal across our many shiftability studies is to build a pharmacological repository of how different drugs ‘shift’ brain processing mechanisms across multiple levels of organisation in autistic and non-autistic individuals.

## 2. Measuring ‘shift’

There is currently no reliable ‘biomarker’ that separates people with an autism diagnosis from controls^12^. For example, functional magnetic resonance imaging (fMRI) data from the EU-AIMS consortium indicates that there is no statistically significant difference in brain activation during facial emotion processing in autism^13^. This work emphasises the message from a recent large-scale meta-analysis carried out by Mottron and colleagues. They found that, even if there are statistically significant case-control differences, the effect sizes reported from ASD case-control analyses are modest at best (the largest is around a standard deviation of 1.0 for Theory of Mind studies^12^) and have gotten smaller over time, despite ever larger study sample sizes^14^. One interpretation of this pattern is that we have too broad a diagnostic definition of autism; another is that the larger the sample size the more likely it is to revert towards the mean; and yet another is that we need to identify more biologically homogeneous subgroups (i.e. stratify). None of these approaches have significantly advanced the field. We propose a different strategy – that measures should be sensitive to individual diversity and capture more ‘foundational’ aspects of brain function; and that they should permit examination of how these functions ‘shift*’* in response to a change in neurochemical pathways in autistic and non-autistic people.

In selecting measures that capture ‘shift’, we must recognise that the genetic and environmental influences which increase the likelihood of neuropsychiatric conditions do not map directly to autism. Instead, they generate the neurobiological conditions for autism and other neurodevelopmental, neurological and mental health conditions, such as schizophrenia, attention deficit hyperactivity disorder, mood disorders, epilepsy and learning difficulties. These are life factors which act on the brain systems developing early, namely sensory systems and emerging interconnecting whole-brain networks^15–20^. Altering these ‘foundational’ aspects of brain development may have cascading effects as the brain matures, generating diversity in cognition and behaviour^21,22^. Secondary and compensatory mechanisms will also continue to act across the lifespan leading to heterogeneous clinical outcomes^23^. However, early changes to the foundational properties of the brain should persist and could therefore be a measurement target of differences in brain function.

In support of this principle, we and others have evidence that the regulation of sensory processing and the functional connectivity of brain networks is associated with ASD across the lifespan. For example, using EEG, Kolenik and colleagues have reported that infants later diagnosed with autism do not suppress neural responses to repeated auditory stimulation^24^; similarly, Piccardi and colleagues identified that infants with elevated likelihood for autism show reduced tactile sensory gating^25^. Using MRI, we have reported that the local functional connectivity of sensory systems is altered in newborns with a higher likelihood of developing autism^15^; and the global functional connectivity of brain networks at full-term predicts emergence of later autistic traits^26^. Moreover, we have reported similar profiles in autistic adults, namely reduced sensory suppression detected using EEG^10^ and altered functional connectivity of sensory systems and whole-brain networks using MRI^27^. This is important because these measures not only capture brain function across age groups, they may also be back-translated to animal models to support a translational bridge^28,29^.

Also important is that selected measures of altered brain function are likely to be sensitive to modulation with a pharmacological challenge. Using fMRI, we have consistently found this to be the case. For example, we have reported that fMRI response can be modulated with single dose drug challenges which target components of the serotonin system (citalopram and tianeptine)^2,3,8,9^, glutamate-GABA system (riluzole)^11^ and endocannabinoid system (cannabidiol)^4,5^. We have also used EEG recordings during sensory stimulation to capture response to GABA_B_ receptor agonist, arbaclofen. We have been able to quantify individual ‘shifts’ in these measures in response to drug challenge and differences at group-level between autistic and non-autistic participants^10,30^. The range of measures used to capture ‘shift’ and their target level of brain organisation is shown in Figure 1.

**Figure 1.**
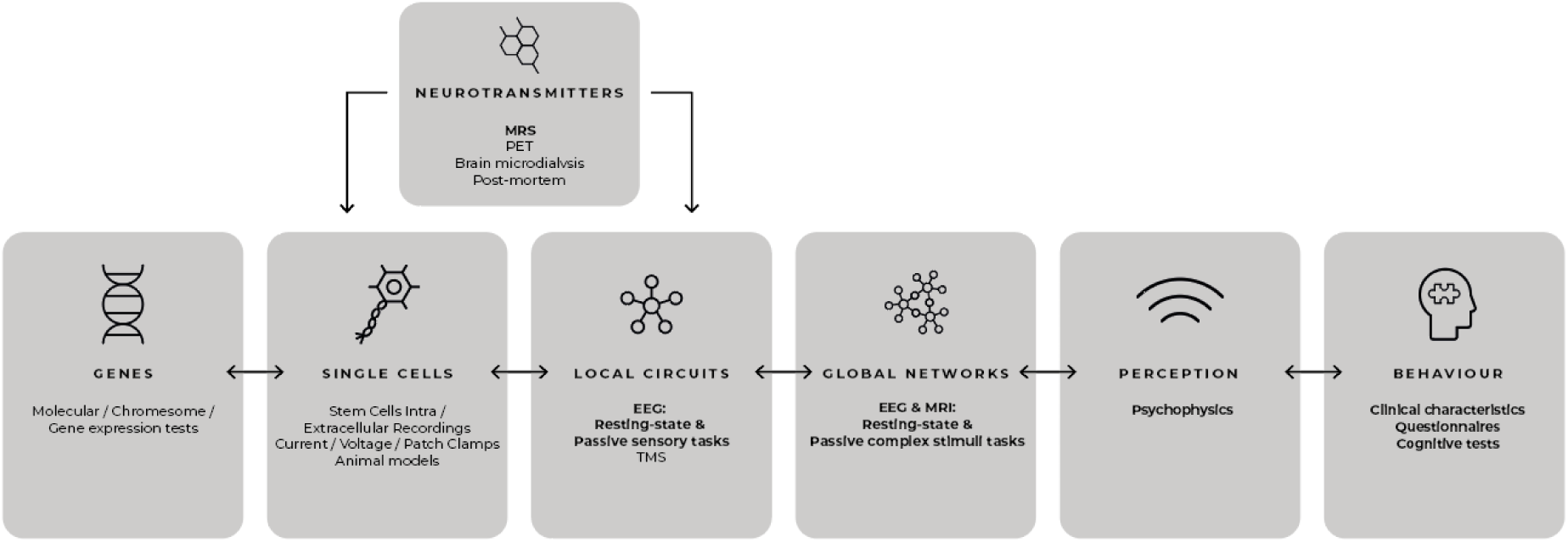
The organisational levels of information processing in the brain and methodologies that probe each level. Methodologies used to detect ‘shift’ that are included in our “shiftability” paradigm and discussed here are shown in bold. Bidirectional arrows represent interaction between organisation levels (adapted from Ahmad & Ellis, 2022^31^).

**Figure 2.**
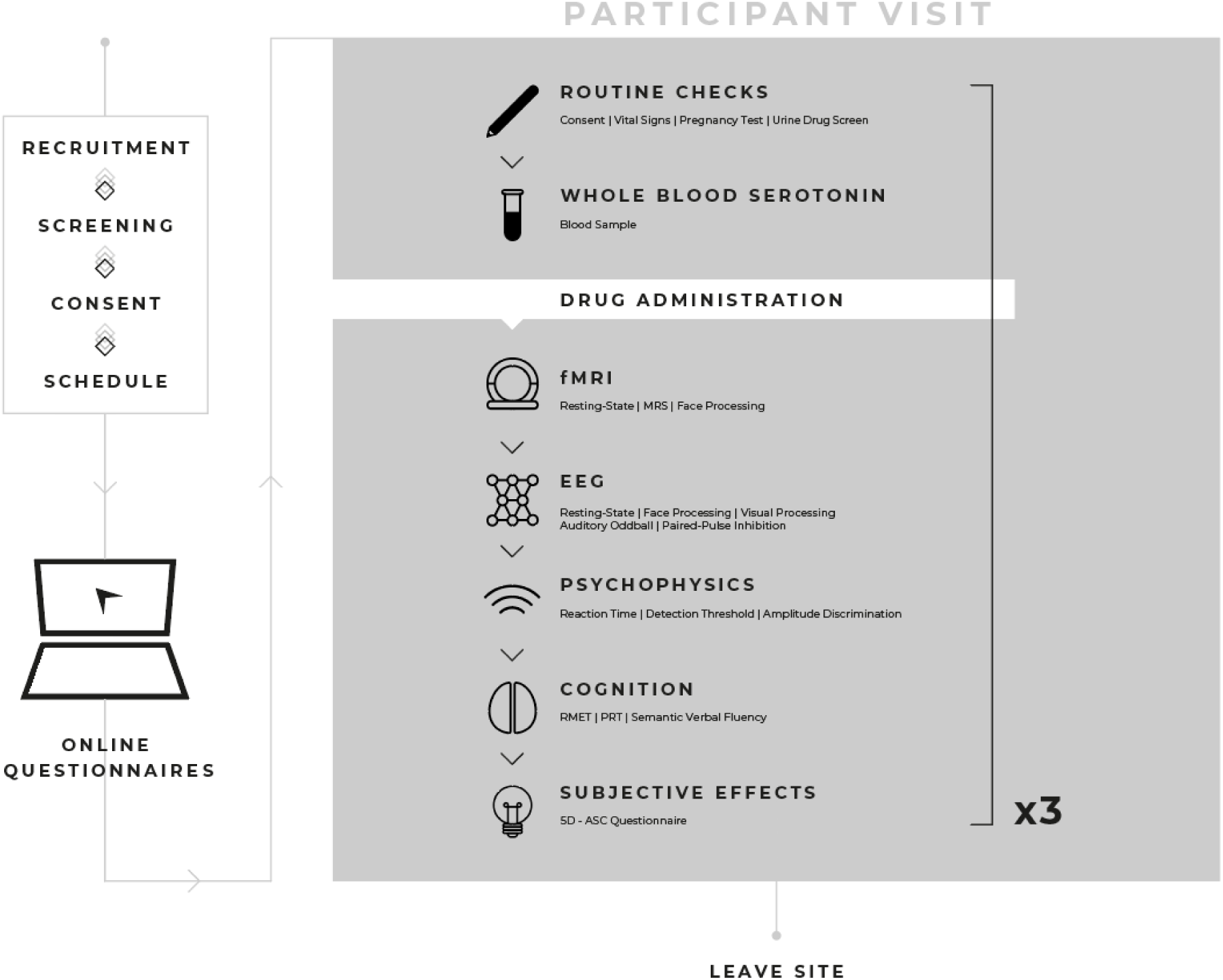
“PSILAUT” recruitment and study procedures. Autistic and non-autistic participants will be recruited from existing local research databases, advertising on the King’s College London website and wider dissemination of study information. Participants are welcome to self-refer. Autistic participants will also be recruited from clinical contacts South London and Maudsley NHS trust, local and national support groups and via a database managed by our collaborators at the Autism Research Centre, University of Cambridge. Interested participants will be sent an information sheet and screened via video call (eg. Zoom) or phone for eligibility. Written consent will be sought after inclusion criteria are confirmed and the participant is then assigned to a study schedule. Participants will be provided with log in details to an online platform (Delosis Ltd., London) to complete a battery of questionnaires remotely. Participants will visit the study site on three separate occasions. A blood sample will be collected on one of the three visits for quantification of whole blood serotonin levels. Each participant will complete an MRI scan session to acquire a structural, resting-state functional MRI scan and a face emotion processing task. Participants will wear a wrist wearable (ActiGraph, Pensacola, FL, USA) following the MRI scan session for the remainder of the study visit. The EEG paradigm will include resting-state and functional activation during a face processing, auditory oddball, visual processing and tactile paired-pulse inhibition tasks. Psychophysical tasks will be collected prior to a cognitive battery which will include the ‘reading the mind in the eyes’ (RMET), probabilistic reversal learning (PRT), both of which will be delivered using PsyTools (Delosis Ltd., London) and a semantic verbal fluency task. The 5-dimensional altered states of consciousness (5D-ASC) questionnaire will then be completed to quantify any subjective effects experienced by participants.

## 3. Targeting the serotonin system in autism using psilocybin

Alterations in the serotonin system are consistently reported in autism and hence, whether there are functional differences in this system is an important focus for our “shiftability” studies. However, the evidence linking serotonin to autism to date, has mostly been indirect from associations and/or correlational studies. For example, polymorphisms in genes for serotonin synthesis, transporters and receptors are associated with autism^32,33^. Elevated whole blood serotonin levels are also reported in one-third of autistic individuals^34,35^. In contrast to these indirect approaches, direct alteration of serotonergic system function *in vivo* can be achieved using pharmacological probes. In animal model systems, this modulation of receptor targets of serotonin has included agonism of 5HT_1A_ and 5HT_7_ receptors to increase social behaviour and reduce stereotypy^36^; and activation of 5HT_2A,_ 5HT_1A_ and 5HT_7_ receptors to shift glutamate-GABA indices of excitation/inhibition (E/I) balance^37,38^, proposed to be altered in autism^39^. In humans, this approach is less frequently taken, but our team has previously reported that acutely elevating serotonin levels with a single dose of selective-serotonin reuptake inhibitor (SSRI) citalopram produces sustained activation of brain regions associated with facial expression processing in autistic adults, but not in controls^2^. This finding suggests that acute non-selective activation of the serotonin system may influence behaviours associated with autism.

There is also evidence implicating specific serotonin receptors in autism. The 5HT_2A_ receptor, encoded by the HT2RA gene, is a robust functional candidate gene in autism^40–43^, and lower cortical 5HT_2A_ receptor binding has been reported to correlate with social communication differences in autism^44^. The 5HT_2A_ receptor is expressed throughout the cortex but especially in regions related to sensorimotor integration^45^ and the so-called default mode network responsible for “self” and “other” processing^46^. At the circuit level, 5HT_2A_ receptor signalling is thought to enhance neural plasticity^47^ and increases cortical glutamate and thalamic GABA levels^38^. However, it is not known whether there are autistic differences in 5HT_2A_ regulation of sensorimotor processes, networks such as the default mode network, and regulation of glutamate-GABA levels is not known. To assess this, we need targeted pharmacological probes. Unlike animal studies, the choice of probe in humans is constrained by the safety and side effect profile of candidate compounds, and few (if any) neuropsychiatric drugs used in people are entirely selective. With this caveat in mind, we selected psilocybin (4-phosphoryloxy-*N.N*-dimethyltryptamine) as a pharmacological probe of the serotonin system based on its affinity for 5HT_2A_Rs.

Psilocybin is a classic psychedelic compound produced by several species of mushrooms, including so-called “magic mushrooms”. Psilocybin is rapidly metabolised into its active component psilocin^48^. Psilocin is a 5HT_2A_R agonist but also binds several serotonin receptors, including 5HT_7_, 5HT_2B_, 5HT_1D_, 5HT_6_, 5HT_5_, 5HT_2C_ & 5HT_1B_ receptors in decreasing order of reported affinity^49^.

Prior studies have used relatively high doses of psilocybin to explore the effects of psychedelics on the brain. However, we will use lower doses (2 mg and 5 mg) in our “shiftability” protocol to assess whether 5HT_2A_R-mediated brain responses are uncomplicated by marked psychedelic experiences. We are confident that our dose range will generate a ‘shift’ in brain function based on evidence using positron emission topography that similar doses of psilocybin engage 5HT_2A_R receptors^50^ and low dose psilocybin is sufficient to alter cognition and obsessive-compulsive behaviour^51,52^. Low doses of the serotonergic psychedelic lysergic acid diethylamide (LSD) and psilocybin-containing mushrooms also acutely alter brain resting-state fMRI and EEG indices in the non-autistic population^53,54^, as well as neural responses to sensory stimuli^55^. Therefore, we expect that 2 mg and 5 mg of psilocybin will expose functional differences in the serotonin system targeted by psilocybin in autistic and non-autistic individuals.

## 4. The “shiftability” paradigm: An example study protocol

### 4.1. Overall design of the ‘PSILAUT’ study

This study will be conducted in accordance with the Declaration of Helsinki at the Institute of Psychiatry, Psychology and Neuroscience (IoPPN) at De Crespigny Park, SE5 8AF, London, United Kingdom. Our study does not address safety or clinical efficacy and the UK Medicines and Health Regulatory Authority (MHRA) has confirmed that our protocol is therefore not a clinical trial of an Investigational Medicinal Product (IMP) as defined by the EU Directive 2001/20/EC. Nevertheless, as our design incorporates a ‘drug intervention’ our protocol has been registered on clinicaltrials.gov for transparency (NCT05651126). Ethical approval has been received from the UK Health Research Authority following review by the Dulwich Research Ethics Committee (reference: 21/LO/0795).

We aim to recruit up to 70 healthy adult participants including 40 autistic and 30 non-autistic control participants in our placebo-controlled, randomised, double-blind, repeated-measures, cross-over, case-control study. All participants will provide written informed consent. Participants will receive either placebo or one of two single doses (2 mg or 5 mg) of oral synthetic COMP360 psilocybin on three separate visits. The order of administration of placebo and psilocybin will be pseudo-randomised to ensure balanced numbers of individuals have placebo or drug on their first visit, and to control for order effects. However, the lowest dose of psilocybin will always precede the higher dose to allow for unblinding should a participant experience unwanted side effects on either of the first two visits. If that visit is a low dose psilocybin visit, we will avoid exposing that participant to the higher dose on a subsequent visit.

The study is an Investigator-Initiated Study sponsored by King’s College London and co-Sponsored by South London and Maudsley NHS Foundation Trust. It is part funded by COMPASS Pathfinder Ltd with infrastructure support from the NIHR-Maudsley Biomedical Research Centre at South London and Maudsley NHS Foundation Trust and King’s College London. COMPASS Pathfinder Ltd are donating psilocybin (as “COMP360”).

### 4.2. ‘PSILAUT’ Protocol Measures

#### Baseline characterisation

A comprehensive baseline characterisation will be obtained. An expert clinical diagnosis of ASD from a recognised UK assessment service will be accepted. This may be supported by the Autism Diagnostic Interview-Revised^56^ where an appropriate informant is available. An Autism Diagnostic Observation Schedule^57^ will be used to assess current symptom level, but if it has already been used to inform the diagnostic assessment in adulthood, it will not be repeated. Additional baseline questionnaires will quantify core autistic features (e.g. social behaviour or sensory differences), relevant cognitive domains (e.g. intolerance of uncertainty and behavioural flexibility) and the symptomology of co-occurring psychiatric conditions.

#### 4.2.1. Neurometabolites

##### Magnetic Resonance Spectroscopy (MRS)

An MRS Hadamard Encoding and Reconstruction of MEGA-Edited Spectroscopy (HERMES) sequence^58^ will be collected during the MRI scan for the dorsal medial prefrontal cortex region. HERMES permits the quantification of levels of metabolites in the living brain and is focused on estimating GABA and Glutamate-glutamine markers of E/I balance. For the purposes of our study, given the evidence that E-I pathways are modulated by 5HT_2A_ receptor action in animal models^38^, we will be able to examine the impact of psilocybin on these tissue level measures of E-I balance.

#### 4.2.2. Local Circuits

##### EEG

###### Resting-state

High-density (64-channel) EEG data will be collected during the resting-state, metrics from which local circuit activity can be derived such as beta and gamma band power/frequency^31^. Oscillations in the beta frequency band at rest, for example, are associated with inhibitory neurotransmitter levels in sensorimotor cortex^59^. This, and other EEG-derived metrics such as aperiodic activity, are considered a proxy measure for E/I balance *in vivo*^31^. Hence, we will be able to examine the impact of psilocybin on these dynamic measures of E-I balance.

##### Passive sensory tasks

###### Visual domain

A visual processing task (contrast saturation) in which steady-state evoked potentials (SSVEPs) are elicited by passive surround suppression stimuli will be conducted. We have shown that SSVEPs during this task are altered in autism^10^. 5HT_2A_Rs are particularly highly expressed in the primary visual cortex^46^, and their agonism alters visual response amplitudes and surround suppression in mouse primary visual cortex^60^. In humans, we expect visual processing to be altered by 5HT_2A_R activation given that the marked visual perceptual changes robustly induced with higher doses of psychedelics are blocked by pretreatment with the 5HT_2_ receptor antagonist, ketanserin^61^.

###### Auditory domain

A conventional auditory oddball paradigm (mismatch negativity, MMN)^62^ will be used to passively measure ‘repetition suppression’ (or habituation) to repetitive auditory stimuli and response to an unexpected ‘deviant’ stimulus (the event-related mismatch negativity MMN response). We and others have observed less repetition suppression in both eight-month-old infants who go on to receive a diagnosis or autism, and adults with a diagnosis of autism^24,30^. Thus, this signal appears linked to autism across infancy to maturity. The impact of autism on the event-related MMN is less consistent and varies with age^63–65^. The latter may in part be due to differences in the serotonin system, as the MMN response can be modulated by acute elevation of serotonin levels by the highly selective SSRI escitalopram^66^. In this study we will test the prediction that psilocybin alters both sensory suppression and MMN in autism differently compared to controls.

###### Tactile domain

A tactile paired-pulse inhibition task will be used to passively investigate EEG responses as objective markers of sensory gating. Aligning with findings in the auditory and visual domain, less tactile neural repetition suppression in this task is associated with autism^25^. As processing of tactile stimuli is also known to be perturbed by 5HT_2A_R agonism with psilocybin^67^, we also expect to elicited functional differences following psilocybin in autistic and non-autistic individuals in this paradigm.

#### 4.2.3. Global Networks

##### Resting-State

###### EEG

Oscillatory power will be assessed across multiple frequency bands during the resting-state. This will include electrodes over key brain regions implicated in autism such as those belonging to the default mode network^68^. Reduced oscillatory power over DMN regions using electrophysiological approaches following 5HT_2A_R activation by psilocybin has been reported previously^69^. Functional connectivity analyses (e.g. within and between brain networks) can also be derived from EEG, and this will complement connectivity analyses from resting-state fMRI.

###### MRI

Participants will undergo a structural and functional MRI scan. Scans will be acquired on a 3.0 Tesla MR Scanner (General Electric Premier). A fMRI scan with a multiband 4 sequence will be acquired during the resting-state, multiband 4 is preferable for connectivity analyses^70^. In addition, multiband sequences will considerably reduce the repetition time (TR), therefore they have the advantage of allowing dynamic functional connectivity analyses. Autistic differences in both ‘averaged’ functional connectivity and dynamic functional connectivity have been reliably reported across different datasets^27,71^. Functional connectivity of brain networks in neurotypical individuals has also be shown to be acutely modulated by 5HT_2A_R activation^72^. Notably, psilocybin alters dynamic functional connectivity, mediated by 5HT_2A_R agonism^73^. It facilitates state transitions and more temporally diverse brain activity in neurotypical individuals^74^. Our study will be the first to examine the effects of psilocybin on conventional and dynamic functional metrics in autistic individuals.

##### Task-dependent MRI and EEG

###### MRI

fMRI studies of face emotion processing in autism have produced inconsistent results. In the largest study to date, no differences between autistic and non-autistic individuals in fMRI response to facial expressions of emotion were observed^13^. However, we have recently examined the fMRI response to facial expressions of emotion in a social brain network before and after administration of the SSRI, citalopram. We reported that the dynamics of the response to faces is different in autism, in that 5HT reuptake inhibition slows habituation^2^. Consistent with this, blockade of 5HT_2A_Rs causes an ‘opposite’ effect and reduces neural responses to emotional faces during fMRI in neurotypical individuals^75^. Therefore, we expect that psilocybin will alter the dynamics of face emotion processing, but differently in autistic individuals.

###### EEG

Event-related potentials (ERPs) in response to face stimuli will also be assessed during EEG. The N170 component, a neural response present at 170ms following the presentation of facial stimuli and can be modulated by 5HT_2A_R activation^61,76,77^. An altered N170 response is associated with social communication differences in autism and may have utility as a stratification marker that is amenable to support^78^. It is also going to be the first prognostic biomarker for autism (or any neurodevelopmental or psychiatric condition) to be approved by regulatory agencies^78^. The incorporation of this task in our protocol therefore will be an important test of whether an autism biomarker can be modified pharmacologically.

#### 4.2.4. Perception

Psychophysical approaches are structured approaches in which stimulus characteristics are tightly controlled, and they provide robust, objective measures of sensory sensitivity by estimating perceptual metrics^79,80^. Serotonin has been directly implicated in tactile perception, such as in affective touch, by studies using tryptophan depletion (which acutely reduces central serotonin levels) alongside psychophysical approaches^81^. Tactile detection threshold and amplitude discrimination will be assessed, as differences in tactile thresholds have already been reported in autism and are associated with outcomes^82^.

#### 4.2.5. Additional measures

##### Questionnaires

The 5-dimensional altered states of consciousness (5D-ASC) questionnaire^83^ will be completed on each visit following the completion of study procedures to quantify the subjective effects of psilocybin, which are primarily mediated by the 5HT_2A_R^84^.

##### Cognitive Battery

‘Theory of mind’ (ie. cognitive empathy, the ability to understand and take into account the mental state of another individual) can be investigated using the ‘reading the mind in the eyes’ (RMET) task^85^. Other cognitive processes in which differences are observed in autism such as language and executive and reward-related functioning (e.g. flexible choice behaviour) will be assessed with a verbal fluency task and probabilistic reversal learning task, respectively^86,87^.

##### Peripheral biochemistry

Participants will be asked to provide a blood sample on one visit prior to placebo/drug administration (their preference). Whole blood serotonin levels will be determined for each individual, given that elevated levels are present in one-third of individuals with autism^34,35^. This will allow us to explore whether any ‘shift’ in brain function in response to psilocybin depends on overall serotonin ‘tone’ as indexed by proxy.

##### Arousal measures

A wrist wearable will be used to passively collect heart rate data, from which autonomic nervous system function indices of heart rate and heart rate variability, can be derived. Heart rate variability is thought to reflect stress or arousal and is modulated by the serotonin system^88^. These autonomic measures have also been reported to be different in autistic individuals compared to controls^89,90^. Therefore, in this study we will examine the action of psilocybin on these indices of arousal and also consider if arousal interacts with other measurements acquired in the protocol.

### 4.3. Data analyses

The overarching goal of our analyses is to assess whether we see a ‘shift’ by psilocybin in autistic and non-autistic individuals for each modality. Both parametric and nonparametric statistical analyses will be used to test hypotheses that the serotonergic targets of psilocybin functioning differently in autistic individuals. Given the heterogeneity of the autistic population and our prior observations that there is a wide range of pharmacological responses in both autistic and non-autistic individuals, we will calculate individual ‘shift’ for each modality, and what characteristics (eg. clinical scores, questionnaire responses, whole blood serotonin) these are associated with. Although we will generate and analyse data from single modalities, post-hoc we will also explore multimodal metrics (ie. associations between modalities) to understand how ‘shifts’ detected across multiple organisational levels are inter-related.

### 4.4. Power analyses

We will use a within-subject, repeated-measures design with a placebo condition so that each subject is their own control, thus increasing statistical power. Results from our prior neuroimaging studies using pharmacological challenge were successful in detecting group differences in MRI metrics with sample sizes of n = < 20^2–7,9,11^. This implies an effects size (ES, expressed in Cohen’s d) in excess of 1.2. In sensory tasks a sample size of n = 16 per cell is estimated to achieve 80% power to detect a medium effect (0.5) at a = 0.05; this has been achieved even in mixed sex groups of 20 participants or fewer. This reflects the literature in which significant group differences are evident even in mixed sex groups of 20 participants or fewer (e.g. n = 16)^91–93^. Our design relies upon participants attending for repeat test sessions. Thus, there is a chance that participants may ‘drop-out’ and need to be replaced, this is accommodated with our sample size.

### 4.5. Limitations

Our protocol requires active participation despite the passive nature of several of our tasks. For example, participants will wear an EEG cap and will be asked to remain focused on the screen during the presentation of sensory stimuli. This may limit generalisability across ages or to autistic individuals with higher support needs.

Even in our very capable adult cohort, participants can get restless or become distracted. Hence, we have included concurrent eye tracking during EEG to control for the potential confound of participant variability in fixation on the screen. However, many of our tasks require minimal or no response from participants and so they are less likely to be impacted by confounds such as individual cognitive difficulties. We hope that this way, should our indices prove worthy of further investigation and/or incorporation in (for example) clinical trials, they will be more accessible for individuals who may otherwise be excluded from drug development studies.

## 5. Conclusion

Our “shiftability” paradigm aims to determine how different organisational scales of brain function are modulated by neurochemical systems; and identify differences between autistic and non-autistic adults at an individual level. Our most recent shiftability study will use psilocybin to test the hypothesis that the serotonin system targeted by psilocybin is different in autistic and non-autistic people. We believe that the results will expand our understanding of brain biology in neurodivergent and neurotypical (autistic and non-autistic) individuals.

We also hope our work will inform a more personalized medicine approach to autism. Not everyone in our study will respond the same way to psilocybin. By capturing individual biology in both autistic and non-autistic people, this experimental medicine approach may help identify autistic individuals whose serotonin system functions no differently from non-autistic people, and who might therefore not be expected to show a clinical response in a clinical trial. And vice versa; those who respond biologically to psilocybin challenge might ultimately benefit clinically. To date, all clinical trials for the core features of autism have failed. This is in large part because they have included participants based on diagnosis alone and measured outcomes without evaluating mechanisms. In depth pharmacological ‘profiling’ adopting some of the methods described here may help avoid the unnecessary expense and likely failure of clinical trials and facilitate the discovery of novel pharmacological support options for those who would like that choice.

## Data Availability

Not applicable.

## Acknowledgements

The PSILAUT study is an Independent Investigator Study (G.M.M.) funded in part by Compass Pathfinder Ltd. The authors also receive support from EU-AIMS (European Autism Interventions)/EU AIMS-2-TRIALS, an Innovative Medicines Initiative Joint Undertaking under Grant Agreement No. 777394. In addition, this paper represents independent research part funded by the infrastructure of the National Institute for Health Research (NIHR) Mental Health Biomedical Research Centre (BRC) at South London and Maudsley NHS Foundation Trust and King’s College London. The views expressed are those of the author(s) and not necessarily those of the NHS, the NIHR or the Department of Health and Social Care. T.P.W. and K.M. are employees of COMPASS Pathfinder Ltd. and hold shares in the company. N.A.P. has consulted for Deerfield Discovery and Research. D.G.M.M. has consulted for Jaguar Gene Therapy LLC. G.M.M. has received funding for investigator-initiated studies from GW Pharmaceuticals and COMPASS Pathfinder Ltd.. G.M.M. has consulted for Greenwich Biosciences, Inc.

